# Laboratory Biomarkers of COVID-19 Disease Severity and Outcome: Findings from a Developing Country

**DOI:** 10.1101/2020.12.29.20248976

**Authors:** Tigist W. Leulseged, Ishmael S. Hassen, Birhanu T. Ayele, Yakob G. Tsegay, Daniel S. Abebe, Mesay G. Edo, Endalkachew H. Maru, Wuletaw C. Zewde, Lydia K. Naylor, Dejene F. Semane, Menayit T. Deresse, Bereket B. Tezera

## Abstract

**Aim:** To identify laboratory biomarkers that predict disease severity and outcome among COVID-19 patients admitted to the Millennium COVID-19 Care Center in Ethiopia.

**Methods:** A retrospective cohort study was conducted among 429 RT-PCR confirmed COVID- 19 patients who were on follow up from July to October 2020 and with complete clinical and laboratory data. Data was described using frequency tables. Robust Poisson regression model was used to identify predictors of COVID-19 disease severity where adjusted relative risk (RR), P-value and 95% CI for RR were used to test significance and interpretation of results. Binary Logistic regression model was used to assess the presence of statistically significant association between the explanatory variables and COVID-19 disease outcome where adjusted odds ratio, P-value and 95% CI for adjusted odds ratio were used for testing significance and interpretation of results

**Results:** Among the 429 patients studied, 182 (42.4%) had Severe disease at admission and the rest 247 (57.6%) had Non-severe disease (15.6% mild and 42.0% moderate). Regarding disease outcome, 45 (10.5%) died and 384 (89.5%) were discharged alive. Age group (ARR= 1.779, 95% CI= 1.405- 2.252, p-value < 0.0001), Neutrophil to Lymphocyte ratio (NLR) (ARR= 4.769, 95% CI= 2.419 - 9.402 p-value <0.0001), Serum glutamic oxaloacetic transaminase (SGOT) (ARR= 1.358, 95% CI= 1.109- 1.662 p-value=0.003), Sodium (ARR= 1.321, 95% CI= 1.091- 1.600 p-value=0.004) and Potassium (ARR= 1.269, 95% CI= 1.059-1.521 p-value=0.010) were found to be significant predictors of COVID-19 disease severity.

The following factors were significantly associated with COVID-19 disease outcome; age group (AOR= 2.767, 95% CI= 1.099 - 6.067, p-value=0.031), white blood cell count (AOR= 4.253, 95% CI= 1.918 - 9.429, p-value=0.0001) and sodium level (AOR= 3.435, 95% CI= 1.439, 8.198, p-value=0.005).

**Conclusions:** The laboratory markers of NLR of above three, raised SGOT and deranged sodium and potassium levels (both hypo- and hyper-states) were found to be significant predictors of developing severe COVID-19 disease. In addition, deranged values of white blood cell count and sodium levels were significantly associated with worse outcome of the disease. Therefore, assessing and monitoring these laboratory markers at the earliest stage of the disease could have a considerable impact in halting disease progression and death.

## INTRODUCTION

The growing threat due to the COVID-19 pandemic has caused numerous losses to the entire world. In Ethiopia as of October 2, 2020, a total of 120,630 cases were identified with 1 864 deaths reported (1). In the absence of a directed therapy and expanding vaccine service for the disease, working on mechanisms to halt the progression of the disease in order to improve disease outcome is the best strategy in addition to promotion of disease preventive mechanisms. To that aim studies to identify important clinical and laboratory biomarkers that can predict disease severity and outcome have been conducted. So far different clinical, laboratory and radiologic markers that can predict disease severity have been identified with varying results in the face of changing behavior of the disease and geographical disparity. Therefore, understanding predictors of disease severity and outcome are crucial to provide early preventive measures for a better outcome especially in economically developing country setup where intensive care setup might not match the increasing demand in the service.

Different laboratory markers are implicated as an indicator of disease severity, progression and outcome. Deranged cell counts, like anemia, polycythemia, leukopenia and leukocytosis with neutrophil predominance and decreased platelet count are found to be associated with severe disease and worse outcome in hospitalized patients (2-9). Similarly raised liver enzymes and total bilirubin levels were identified in severe and critical patients (6, 10-15).

Raised inflammatory response of the body as manifested by raised laboratory values of various interleukins and C-reactive proteins are also reported (4, 7, 16-18). In addition, raised coagulation markers like fibrinogen and prothrombin time are identified in severe and critical patients (2, 4).

Electrolyte imbalance in both directions, hypo- and hyper- levels were reported for sodium, potassium and calcium levels among patients with severe disease and worse outcome, hypothesized to result from the effect of the disease on the body system or the medication side effects (19-21).

There is limited study that assessed the role of laboratory markers in predicting disease severity and outcome in the African setup. Since the number of infected cases and mortality rate is reported to be lower than those reported in the non-African setup, understanding the disease predictors in our setup is crucial. Therefore, the aim of this study was to identify laboratory biomarkers that predict disease severity and outcome among COVID-19 patients admitted to the Millennium COVID-19 Care Center in Ethiopia from July to October 2020.

## METHODS AND MATERIALS

### Study setting, design and population

An institution based retrospective cohort study was conducted at Millennium COVID-19 Care Center (MCCC), a makeshift hospital in Addis Ababa, Ethiopia. The follow up was made from July to October, 2020. The source population was all cases of COVID-19 admitted at MCCC with a confirmed diagnosis of COVID-19 using RT-PCR, as reported by a laboratory given mandate to test such patients by the Ethiopian Federal Ministry of Health and who were on follow up from July to October, 2020 (22).

All consecutively admitted Severe COVID-19 patients during the follow up period with complete clinical and laboratory data were included in the study. With these criteria, a total of 429 COVID-19 patients were included in the final analysis.

### Eligibility criteria

All COVID-19 patients who were on treatment and follow up at the center from July to October, 2020 and with complete baseline clinical and laboratory data were included.

### Data Collection Procedures and Quality Assurance

Data was extracted from patients’ admission, follow up and discharge charts using a pretested electronic data abstraction tool that is adopted from the WHO CRF form. Data extractors were trained on the tool and appropriate infection prevention and control measures were followed.

Data consistency and completeness was checked before an attempt was made to enter the code and analyze the data.

### Data Management and Data Analysis

Data was summarized using frequency tables and percentages.

To identify predictors of COVID-19 disease severity, Robust Poisson regression model was used. Variables significantly associated with disease severity at 25% level of significance in the Univariate analysis were were considered in the multivariable model. In the final model; adjusted RR, P-value and 95% CI for RR were used to test significance and interpretation of results. Variables with p-value ≤ 0.05 were considered as significant predictors of disease severity.

To assess the presence of statistically significant association between COVID-19 disease outcome (death) and the explanatory variables, Binary Logistic regression model was used. Univariate analysis was done to screen out independent variables to be used in the multivariable Binary Logistic regression model at 25% level of significance. The adequacy of the final model was assessed using the Hosmer and Lemeshow goodness of fit test and the final model fitted the data well p-value = 0.876). For the Binary Logistic regression, 95% confidence interval for AOR was calculated and variables with p-value ≤ 0.05 were considered as statistically associated with disease outcome.

All analyses were performed using STATA software version 14 (College Station, TX).

## RESULT

### COVID-19 Severity and Disease Outcome

Among the 429 patients studied, 182 (42.4%) had severe disease and the rest 247 (57.6%) had non-severe disease (15.6% mild and 42.0% moderate) at admission. Regarding disease outcome, 45 (10.5%) died and 384 (89.5%) were discharged alive.

### Socio-demographic, comorbid illness and presenting symptoms

More than half of the participants were younger than 50 years (60.8%) and males (64.1%). One hundred eighty eight (43.8%) had a history of one or more preexisting co-morbid illness. The majority had hypertension (25.2%), Type II diabetes mellitus (TIIDM) (19.8%), Asthma (6.1%) and cardiac disease (5.6%). Other co-morbid illness including chronic diseases of the lung, kidney, liver and neurology constituted less than 1% of the total cases.

More than three fourth (79.0%) of the patients were symptomatic at presentation. The commonest symptoms were cough (69.2%), followed by fatigue (29.1%) and fever (26.1%). (**Table 1**)

**Table 1:**
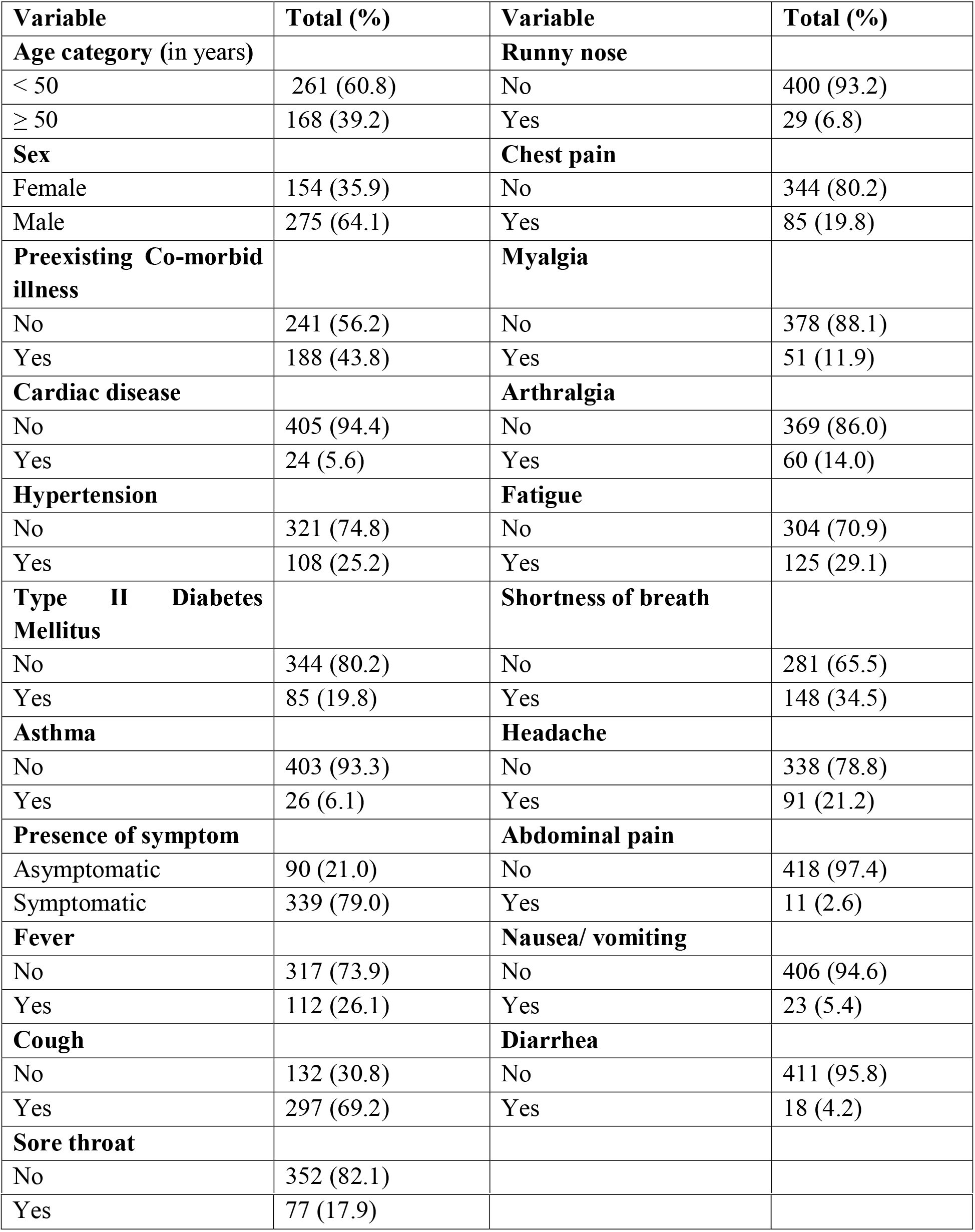
Socio–demographic, co-morbid illness and presenting symptom related variables among COVID-19 patients (n=429)

| **Variable** | **Total (%)** | **Variable** | **Total (%)** |
| --- | --- | --- | --- |
| **Age category (in years)** |  | **Runny nose** |  |
| < 50 | 261 (60.8) | No | 400 (93.2) |
| ≥ 50 | 168 (39.2) | Yes | 29 (6.8) |
| **Sex** |  | **Chest pain** |  |
| Female | 154 (35.9) | No | 344 (80.2) |
| Male | 275 (64.1) | Yes | 85 (19.8) |
| **Preexisting Co-morbid illness** |  | **Myalgia** |  |
| No | 241 (56.2) | No | 378 (88.1) |
| Yes | 188 (43.8) | Yes | 51 (11.9) |
| **Cardiac disease** |  | **Arthralgia** |  |
| No | 405 (94.4) | No | 369 (86.0) |
| Yes | 24 (5.6) | Yes | 60 (14.0) |
| **Hypertension** |  | **Fatigue** |  |
| No | 321 (74.8) | No | 304 (70.9) |
| Yes | 108 (25.2) | Yes | 125 (29.1) |
| **Type II Diabetes Mellitus** |  | **Shortness of breath** |  |
| No | 344 (80.2) | No | 281 (65.5) |
| Yes | 85 (19.8) | Yes | 148 (34.5) |
| **Asthma** |  | **Headache** |  |
| No | 403 (93.3) | No | 338 (78.8) |
| Yes | 26 (6.1) | Yes | 91 (21.2) |
| **Presence of symptom** |  | **Abdominal pain** |  |
| Asymptomatic | 90 (21.0) | No | 418 (97.4) |
| Symptomatic | 339 (79.0) | Yes | 11 (2.6) |
| **Fever** |  | **Nausea/ vomiting** |  |
| No | 317 (73.9) | No | 406 (94.6) |
| Yes | 112 (26.1) | Yes | 23 (5.4) |
| **Cough** |  | **Diarrhea** |  |
| No | 132 (30.8) | No | 411 (95.8) |
| Yes | 297 (69.2) | Yes | 18 (4.2) |
| **Sore throat** |  |  |  |
| No | 352 (82.1) |  |  |
| Yes | 77 (17.9) |  |  |

### Baseline Laboratory biomarkers

The complete blood count of the study participants showed that more than one-third (36.1%) had polycythemia and only 23 (5.4%) were anemic. Close to one-third had deranged white blood cell count (WBC), with 61 (14.2%) had leukopenic and 69 (16.1%) had leukocytosis. Majority had neutrophil predominant (66.2%) and lymphopenic cell count profile (71.3%). Only a quarter (26.8%) of patients had a normal Neutrophil to Lymphocyte ratio (NLR) of ≤ 3. Majority (85.1%) of the patients had a normal platelet count.

Three hundred forty seven (80.9%) of the patients had a raised urea value and 62 (14.5%) had raised creatinine. Raised liver enzymes level of Serum glutamic pyruvic transaminase (SGPT), Serum glutamic oxaloacetic transaminase (SGOT) and Alkaline phosphatase (ALP) were recorded on 43.6%, 24.5% and 18.2% of the patients, respectively.

Abnormal serum levels of Sodium (Na) and Potassium (K) were observed in considerable proportion of patients. A higher proportion of deranged value was observed for hyponatremia and hyperkalemia which were observed in 63 (14.7%) and 127 (29.6%) of the patients, respectively. **(Table 2**)

**Table 2:** Baseline Laboratory biomarkers related variables among COVID-19 patients (n=429)

| Variables | Total (%) | Variables | Total (%) |
| --- | --- | --- | --- |
| **Hematocrit (%)** |  | **Urea (mg/dl)** |  |
| <36 | 23 (5.4) | < 20 | 82 (19.1) |
| 36-45 | 251 (58.5) | $\geq 20$ | 347 (80.9) |
| >45 | 155 (36.1) |  |  |
| **White Blood Cell (cells/ul)** |  | **Creatinine (mg/dl)** |  |
| <4.5 | 61 (14.2) | <0.6 | 48 (11.2) |
| 4.5-11 | 299 (69.7) | 0.6-1.1 | 319 (74.4) |
| >11 | 69 (16.1) | >1.1 | 62 (14.5) |
| **Neutrophil %** |  | **SGPT (IU/L)** |  |
| <40 | 21 (4.9) | < 41 | 242 (56.4) |
| 40-70 | 124 (28.9) | $\geq 41$ | 187 (43.6) |
| >70 | 284 (66.2) | **SGOT (IU/L)** |  |
| **Lymphocyte %** |  | < 40 | 324 (75.5) |
| <20 | 306 (71.3) | $\geq 40$ | 105 (24.5) |
| 20-50 | 119 (27.7) | **ALP (IU/L)** |  |
| >50 | 4 (0.9) | < 100 | 351 (81.8) |
| **NLR** |  | ≥ 100 | 78 (18.2) |
| ≤ 3 | 115 (26.8) | **Na** (mequ/l) |  |
| >3 | 314 (73.2) | <135 | 63 (14.7) |
| **Platelet count** (cells/ul) |  | 135-145 | 349 (81.4) |
| <150 | 36 (8.4) | >145 | 17 (4.0) |
| 15-450 | 365 (85.1) | **K** (mequ/l) |  |
| >450 | 28 (6.5) | <3.5 | 17 (4.0) |
|  |  | 3.5-4.5 | 285 (66.4) |
|  |  | >4.5 | 127 (29.6) |

### Predictors of COVID-19 disease severity (Severe Vs Non-Severe)

Univariate analysis at 25% level of significance was conducted and age group, sex, hypertension, type II diabetes mellitus, fever, sore throat, myalgia, arthralgia, fatigue, headache, hematocrit (Hct), WBC, platelet count, NLR, urea, creatinine, SGPT, SGOT, ALP, Na and K were found to be predictors of COVID-19 disease severity.

On the multivariable Robust Poisson Regression, after adjusting for other covariates, age group, fever, fatigue, NLR, SGOT, Na and K were found to be significant predictors of COVID-19 disease severity at 5% level of significance.

After adjusting for other covariates, being 50 years and older increased risk of developing severe disease by 1.779 (ARR= 1.779, 95% CI= 1.405-2.252, p-value < 0.0001).

The presenting symptoms of patients particularly fever and fatigue were found to be significant predictors of disease severity showing a 1.252 (ARR= 1.252, 95% CI= 1.019-1.536, p-value=0.032) and 1.414 (ARR= 1.414, 95% CI= 1.153-1.732 p-value=0.001) times increased risk of having severe disease as compared to patients with no such symptoms, respectively.

NLR of greater than 3 was associated with a 4.769 times increased risk of developing severe disease as compared with those with a value of 3 and lower (ARR= 4.769, 95% CI= 2.419, 9.402 p-value <0.0001).

Having a raised SGOT of 41 and above increased the risk of having severe disease by 35.8% compared to those with value of a normal range (ARR= 1.358, 95% CI= 1.109, 1.662 p-value=0.003).

The risk of developing severe disease among patients with deranged values of Na and K level (both hypo- and hyper- levels) was 1.321 (ARR= 1.321, 95% CI= 1.091, 1.600 p-value=0.004) and 1.269 (ARR= 1.269, 95% CI= 1.059, 1.521 p-value=0.010) times that of patients with a normal values of Na and K, respectively. (**Table 3**)

**Table 3:** Results of the final multivariable Robust Poisson Regression model among COVID-19 patients (n=429)

| Variable | CRR(95% CI) | ARR(95% CI) | p-value |
| --- | --- | --- | --- |
| **Age group** ($\geq 50$ Vs $< 50$ Years) | 2.544 (2.024, 3.197) | **1.779 (1.405, 2.252)** | **<0.0001\*** |
| **Sex** (Male Vs Female) | 1.363 (1.059, 1.753) | 1.204 (0.983, 1.474) | 0.072 |
| **Hypertension** (Yes Vs No) | 1.498 (1.206, 1.862) | 1.117 (0.914, 1.364) | 0.277 |
| **Type II Diabetes Mellitus** (Yes Vs No) | 1.576 (1.264, 1.963) | 0.989 (0.789, 1.236) | 0.916 |
| **Fever** (Yes Vs No) | 1.572 (1.270, 1.947) | **1.252 (1.019, 1.536)** | **0.032\*** |
| **Sorethroat** (Yes Vs No) | 1.206 (0.931, 1.563) | 1.169 (0.929, 1.470) | 0.184 |
| **Myalgia** (Yes Vs No) | 1.581 (1.237, 2.021) | 0.877 (0.695, 1.106) | 0.268 |
| **Arthralgia** (Yes Vs No) | 1.732 (1.389, 2.160) | 1.213 (0.948, 1.551) | 0.124 |
| **Fatigue** (Yes Vs No) | 2.342 (1.985, 2.979) | **1.414 (1.153, 1.732)** | **0.001\*** |
| **Headache** (Yes Vs No) | 1.330 (1.052, 1.683) | 1.085 (0.879, 1.338) | 0.448 |
| **Hematocrit** ($>45$ Vs $\leq 45\%$) | 0.766 (0.597, 0.981) | 0.889 (0.728, 1.088) | 0.254 |
| **White blood cell count** ($<4.5/>11/\text{ul}$ Vs $4.5-11 \times 10^3/\text{ul}$) | 1.247 (0.997, 1.561) | 0.958 (0.796, 1.152) | 0.646 |
| **NLR** ($>3$ Vs $\leq 3$) | 7.966 (4.049, 15.669) | **4.769 (2.419, 9.402)** | **<0.0001\*** |
| **Platelet Count** ($<150/>450/\text{ul}$ Vs $150-450 \times 10^3/\text{ul}$) | 1.555 (1.230, 1.966) | 1.238 (0.984, 1.557) | 0.069 |
| **Urea** ($\geq 20$ Vs $<20$ mg/dl) | 1.197 (0.879, 1.631) | 1.122 (0.852, 1.477) | 0.413 |
| **Creatinine** ($<0.6/>1.1$ Vs $0.6-1.1$ mg/dl mg/dl) | 1.256 (0.997, 1.582) | 1.009 (0.809, 1.258) | 0.936 |
| **SGPT** ($\geq 41$ Vs $<41$ IU/L) | 1.509 (1.211, 1.882) | 1.056 (0.873, 1.277) | 0.576 |
| **SGOT** ($\geq 40$ Vs $<40$ IU/L) | 2.420 (1.994, 2.936) | **1.358 (1.109, 1.662)** | **0.003\*** |
| **ALP** ($\geq 100$ Vs $<100$ IU/L) | 1.612 (1.292, 2.009) | 1.021 (0.846, 1.232) | 0.828 |
| **Na** ($<135/>145$ mequ/l Vs $135-145$ mequ/l) | 2.092 (1.724, 2.539) | **1.321 (1.091, 1.600)** | **0.004\*** |
| **K** ($<3.5/>4.5$ mequ/l Vs $3.5-4.5$ mequ/l) | 1.518 (1.225, 1.881) | **1.269 (1.059, 1.521)** | **0.010\*** |

**Note:** CRR, Crude Risk ratio; ARR, Adjusted Risk ratio; CI, Confidence interval; \*statistically significant

### Predictors of COVID-19 Disease Outcome (Death Vs Recovery)

Crude analysis of each independent variable with disease outcome was run at 25% level of significance. From univariate analysis; age group, sex, hypertension, TIIDM, sore throat, chest pain, myalgia, arthralgia, fatigue, respiratory rate, Hct, WBC, platelet count, urea, creatinine, SGPT, SGOT, ALP, Na and K were found to be significantly associated with COVID-19 disease outcome.

However; only age group, WBC and Na level were found to be significantly associated with disease outcome in the multivariable binary logistic regression model at 5% level of significance.

Accordingly, after adjusting for other covariates, the odds of dying among patients who were 50 years and older was 2.767 times compared with those less than 50 years of age (AOR= 2.767, 95% CI= 1.099, 6.067, p-value=0.031).

The laboratory markers that are found to be significant determinants of disease outcome were WBC count and Na level. After being adjusted for other factors, having deranged laboratory markers (both lower and raised values) of WBC count and Na level were associated with 4.253 times (AOR= 4.253, 95% CI= 1.918, 9.429, p-value=0.0001) and 3.435 times (AOR= 3.435, 95% CI= 1.439, 8.198, p-value=0.005) higher odds of dying compared to those with normal values for these markers. (**Table 4**)

**Table 4:** Result of Multivariable Binary logistic regression model among COVID-19 patients (n=429)

| Variable | COR(95% CI) | AOR(95% CI) | p-value |
| --- | --- | --- | --- |
| **Age category** ($\geq 50$ Vs $< 50$ Years) | 5.072 (2.536, 10.144) | **2.767 (1.099, 6.967)** | **0.031\*** |
| **Sex** (Male Vs Female) | 1.834 (0.901, 3.733) | 2.076 (0.841, 5.121) | 0.113 |
| **Hypertension** (Yes Vs No) | 2.691 (1.427, 5.074) | 1.567 (0.640, 3.836) | 0.325 |
| **Type II Diabetes Mellitus** (Yes Vs No) | 3.926 (2.059, 7.488) | 1.774 (0.718, 4.385) | 0.214 |
| **Sorethroat** (Yes Vs No) | 0.416 (0.144, 1.197) | 0.467 (0.122, 1.786) | 0.266 |
| **Chestpain** (Yes Vs No) | 1.991 (1.007, 3.936) | 1.104 (0.418, 2.917) | 0.841 |
| **Myalgia** (Yes Vs No) | 1.715 (0.750, 3.922) | 0.911 (0.255, 3.253) | 0.886 |
| **Arthralgia** (Yes Vs No) | 1.909 (0.890, 4.093) | 2.441 (0.739, 8.069) | 0.143 |
| **Fatigue** (Yes Vs No) | 2.610 (1.395, 4.883) | 0.965 (0.376, 2.476) | 0.940 |
| **Respiratory rate** ($> 20$ Vs $\leq 20$/min) | 2.665 (1.283, 5.537) | 2.145 (0.853, 5.396) | 0.105 |
| **Hematocrit** ($> 45$ Vs $\leq 45\%$) | 0.539 (0.265, 1.097) | 0.549 (0.229, 1.319) | 0.180 |
| **White blood cell count** ($< 4.5$/$> 11$/ul Vs $4.5$-$11 \times 10^3$/ul) | 4.554 (2.392, 8.669) | **4.253 (1.918, 9.429)** | **0.0001\*** |
| **Platelet Count** ($< 150$/$> 450$/ul Vs $150$-$450 \times 10^3$/ul) | 1.746 (0.817, 3.731) | 0.732 (0.254, 2.111) | 0.563 |
| **Urea** ($\geq 20$ Vs $< 20$ mg/dl) | 2.613 (0.909, 7.514) | 1.916 (0.566, 6.480) | 0.296 |
| **Creatinine** (<0.6/>1.1 Vs 0.6-1.1 mg/dl mg/dl) | 3.217 (1.712, 6.048) | 1.956 (0.829, 4.614) | 0.126 |
| **SGPT** (≥41 Vs <41 IU/L) | 2.892 (1.506, 5.552) | 2.013 (0.840, 4.822) | 0.116 |
| **SGOT** (≥40 Vs <40 IU/L) | 5.283 (2.782, 10.032) | 1.534 (0.644, 3.655) | 0.334 |
| **ALP** (≥100 Vs <100 IU/L) | 4.025 (2.094, 7.737) | 1.825 (0.779, 4.273) | 0.166 |
| **Na** (<135/>145 mequ/l Vs 135-145 mequ/l) | 7.477 (3.889, 14.375) | **3.435 (1.439, 8.198)** | **0.005\*** |
| **K** (<3.5/>4.5 mequ/l Vs 3.5-4.5 mequ/l) | 2.054 (1.102, 3.829) | 1.299 (0.583, 2.897) | 0.522 |

**Note:** COR, Crude Odds ratio; AOR, Adjusted Odds ratio; CI, Confidence interval; \*statistically significant

## DISCUSSION

In this study, we assessed the effect of clinical and laboratory markers on COVID-19 disease severity and outcome among 429 COVID-19 patients who were admitted to Millennium COVID-19 Care Center in Ethiopia from July to October 2020. In this study, we identified the markers of disease severity and outcome. Timely identification of biomarkers of COVID-19 disease severity and death would help to provide targeted intervention and patient management. Among the 429 patients studied, 247 (57.6%) had Non-severe disease (15.6% mild and 42.0% moderate) and the rest 182 (42.4%) had Severe disease at admission. Regarding disease outcome, 45 (10.5%) died and 384 (89.5%) were discharged alive. Age group, NLR, SGOT, Sodium and Potassium were found to be significant predictors of COVID-19 disease severity. In addition, the following factors were significantly associated with the COVID-19 disease outcome; age group, white blood cell count and sodium level.

On the robust Poisson regression model, age group, sex, hypertension, type II diabetes mellitus, fever, sorethroat, myalgia, arthralgia, fatigue, headache, Hct, WBC, NLR, platelet count, urea, creatinine, SGPT, SGOT, ALP, Na and K were found to be significant predictors of COVID-19 disease severity on the crude analysis. But after controlling for the above variables, age group, fever, fatigue, NLR, SGOT, Na and K were found to be predictors of COVID-19 disease severity at 5% level of significance.

Accordingly, after adjusting for other covariates, being 50 years and older was found to be associated with a 1.779 times increased risk of developing severe disease. Age is implicated to be associated with severe disease and outcome in different studies conducted globally (23-26). Similar finding is also reported from studies conducted in our country (27, 28). The reasons behind could be the increased possibility of weaker immune defense mechanism and co-morbid illnesses making older individuals prone to different illnesses with severe progression and worst outcome.

Symptoms of fever and fatigue were found to be significant predictors of disease severity showing a 1.252 and 1.414 times increased risk of having severe disease as compared to patients with no such symptoms, respectively. Having symptomatic disease, other than symptoms used in disease classification, in general is reported to delay disease recovery and also found to be associated with more severe disease category (28, 29).

NLR of greater than three is associated with a 4.769 times increased risk of developing severe disease as compared with those with a value of three and less. As an indicator of the body’s systemic inflammation, a raised level of this inflammatory biomarker above three indicates the body’s stress. With more stress, as in severe and critical patient states, the level increases to even higher level. Therefore, this biomarker is an indirect indication of the body’s stress level due to the severity of the disease. This is also similar with findings from other studies where NLR was found to an important predictor of disease severity and outcome (3, 5-8).

Having a raised SGOT of 41 and above was associated with a 1.358 times increased risk of having severe disease as compared with those with value of a normal range. Different mechanisms are pointed to the possible effect of the SARS-CoV-2 Virus on liver; the virus could directly affect the hepatocytes or the liver could get injured indirectly through enhanced inflammatory response due to the raised immune markers and drug toxicity that are meant to treat or halt the progression of the disease resulting in liver damage and thereby an increase in liver enzymes (30). Raised in liver enzymes associated with more severe disease category is also reported in other studies (10-15).

The risk of developing severe disease among patients with deranged values of Na and K level (both hypo- and hyper- levels) was 1.321 and 1.269 times than patients with a normal values of Na and K, respectively. Electrolyte imbalances in any disease condition can result from fluid losses from the body through different routes, renal damage and effect of medication that is administered to treat the disease and/or concomitant illnesses. It is also implicated to be due to the decreased activity of angiotensin-converting enzyme 2 which is claimed to be a receptor for the severe acute respiratory syndrome coronavirus 2. This is also demonstrated in other studies, where both increased and decreased levels of Na and K are found to be associated with severe disease states (19-21).

On the assessment of factors associated with disease outcome, the univariate binary logistic regression at 25% level of significance shows that age group, sex, hypertension, type II diabetes mellitus, sorethroat, chest pain, myalgia, arthralgia, fatigue, respiratory rate, Hct, WBC, platelet count, Urea, creatinine, SGPT, SGOT, ALP, Na and K were found to be significantly associated with COVID-19 disease outcome. On further multivariable analysis at 5% level of significance, age group, WBC and Na level were found to be significantly associated with disease outcome.

Accordingly, after adjusting for other covariates, the odds of dying among patients who were 50 years and older was 2.767 times compared with those less than 50 years of age. Similar to the above finding where age is associated with more severe disease condition, it is also implicated to be associated with severe disease outcome.

The laboratory markers that are found to be significant determinants of disease outcome were white blood cell count and sodium level.

After being adjusted for other factors, having deranged laboratory markers (both lower and raised values) of white blood cell count was associated with a 4.253 times higher odds of dying compared to those with normal values. Both leukopenia and leukocytosis are found to be associated with severe COVID-19. Having decreased white cell count increases the potential to develop serious infection and expansion of an already existing pathogen thereby leading to the development of critical disease stage in any infection. Similarly, although a raised white cell count is an indication of a strong immune system responding to external threat, it also implies that the body is under a lot of stress from the pathogenic organism.

Similarly, having deranged laboratory markers (both lower and raised values) of Na level was associated with a 3.435 times higher odds of dying compared to those with normal values for this marker. As discussed above, hyponatremia and hypernatremia that can result from different underlying disease conditions or from the medications given to treat the disease are found to be associated with severe disease and worse outcomes as demonstrated by different studies as well (20, 21).

## CONCLUSION

In this study we have assessed laboratory markers that can predict disease severity and outcome. Accordingly, NLR of above three, raised SGOT and deranged Na and K levels (both hypo- and hyper-states) were found to be significant predictors of developing severe COVID-19 disease. In addition, deranged values of WBC and Na levels were significantly associated with worse outcome of the disease.

Therefore, assessing and monitoring these laboratory markers at the earliest stage of the disease could have a considerable input in halting disease progression and death.

## Data Availability

All relevant data are available upon reasonable request.

## Declaration

### Ethical Considerations

The study was conducted after obtaining ethical clearance from St. Paul’s Hospital Millennium Medical College Institutional Review Board. Written informed consent was obtained from the participants. The study had no risk/negative consequence on those who participated in the study. Medical record numbers were used for data collection and personal identifiers were not used in the research report. Access to the collected information was limited to the principal investigator and confidentiality was maintained throughout the project.

### Authors Contribution

TWL conceived and designed the study, revised data extraction sheet, performed statistical analysis, and drafted the initial manuscript. ISH contributed in the conception and design of the study, undertook review and interpretation of the data, revised the manuscript and approved the final version. BTA assisted in the statistical analysis, undertook review and interpretation of the data, revised the manuscript and approved the final version. YGT, DSA, MGD, EHM, WCZ, LKN, DFS, MTD and BBT contributed to the conception, obtained patient data, undertook review and interpretation of the data, revised the manuscript and approved the final version.

## Acknowledgment

The authors would like to thank St. Paul’s Hospital Millennium Medical College for facilitating the research work.

## Availability of data and materials

All relevant data are available upon reasonable request.

